# Age-adjusted trends in the diastolic and systolic heart failure in the United States over recent years based on race and gender with higher trends in Men and African Americans

**DOI:** 10.1101/2025.08.01.25332699

**Authors:** Hannah Kirsch, Mehrtash Hashemzadeh, Mohammad Reza Movahed

**Affiliations:** University of Arizona College of Medicine, Phoenix, AZ; University of Arizona Sarver Heart Center, Tucson, AZ

**Keywords:** heart failure, systolic heart failure, diastolic heart failure, congestive heart failure, CHF, cardiomyopathy, systolic dysfunction, diastolic dysfunction

## Abstract

**Background:** Heart failure (HF) is a leading cause of hospitalization in the United States. The goal of this study was to evaluate contemporary population-level trends and demographic disparities in age-adjusted hospitalization rates for systolic heart failure (SHF) and diastolic heart failure (DHF).

**Methods:** We analyzed discharge data from the National Inpatient Sample (NIS) database, years 2016 to 2020, for adults aged 20 and older. HF subtypes were identified using ICD-10 codes. Age-adjusted hospitalization rates per 100,000 population were calculated and stratified by year, sex, and race.

**Results:** From 2016 to 2020, the age-adjusted DHF hospitalization rate increased from 219.4 (95% CI: 201.4–237.1) to 303.1 (95% CI: 277.7–328.5) per 100,000. SHF rates rose from 211.7 (95% CI: 194.7–228.7) to 262.6 (95% CI: 240.6–284.6). Hospitalizations for SHF were more common in men than women across all years; in 2020, the SHF hospitalization rate in men was 370.6 (95% CI: 323.8-417.4) compared to 171.9 (95% CI: 152.6–191.1) in women. Black patients consistently had the highest SHF and DHF hospitalization rates. In 2020, the DHF rate among Blacks was 418.3 (95% CI: 328.9–507.7) versus 284.8 (95% CI: 255.0–314.6) among Whites, and the SHF rate was 403.6 (317.3–478.8) versus 227.5 (95% CI: 203.7–251.3), respectively.

**Conclusions:** SHF and DHF age-adjusted hospitalization rates are rising significantly, with pronounced disparities by sex and race. Men and Black patients are disproportionately impacted.

## Introduction

Heart failure (HF) is a leading cause of hospitalization in the United States, with recent national estimates nearly reaching or exceeding 1 million hospitalizations per year ^1, 2, 3^— and its prevalence is expected to continue rising in the years to come. ^5, 6, 7^ An aging population overall is thought to be one of the largest contributing factors to this expected rise in HF, ^5, 6^ given that HF incidence does increase significantly with age. ^7, 8, 9, 10^

Presently, HF hospitalizations account for the majority of heart failure-associated healthcare expenditures and are associated with substantial morbidity and mortality. ^1, 2, 11^ As the burden of heart failure continues to rise, the associated strain on healthcare systems and economic resources can be expected to grow substantially, to the potential detriment of patient quality of care. ^1, 5^ Therefore, characterization of recent trends in heart failure hospitalization is of significant interest.

Importantly, marked disparities in HF burden by sex and race have been consistently documented. Overall prevalence of HF is higher in men than in women and SHF is substantially more common in men. ^12, 13, 14, 15, 16^ Racial disparities are also pronounced. Black patients bear a disproportionate burden compared to White patients. ^13, 14, 17^ These patterns have been observed consistently and persist despite advances in therapy.

Despite projections of increasing prevalence, prior analyses of national HF hospitalization trends indicated a steady decline in overall HF hospitalization rates from the early 2000s to 2013. ^18, 19^ More recent analyses of national trends do support a decline in HF hospitalization trends until 2013 but indicate that this trend has since reversed and rates have increased from 2014-2018. ^20, 21^, These findings indicate a dynamic epidemiological landscape and highlight the need for continued surveillance of HF hospitalization trends.

Additionally, systolic and diastolic HF are distinct clinical entities with differing pathophysiological mechanisms and comorbidity profiles. ^10, 16, 19^ Recent epidemiological data has highlighted a shift towards increasing prevalence of diastolic HF compared to systolic HF. ^24, 25, 26, 27, 28^ Prior studies have shown that DHF is more common in women and elderly individuals compared to SHF. ^10, 29, 30, 31, 32, 33^ Understanding subtype-specific trends is important to accurately characterize the evolving burden of HF.

This study addresses the gap in updated national data by examining age-adjusted hospitalization trends for SHF and DHF from 2016 to 2020 using the National Inpatient Sample (NIS). We assess these trends stratified by sex and race to better characterize current demographic disparities.

## Methods

### Data Source

Deidentified, publicly available patient data were obtained from the National Inpatient Sample (NIS). The NIS database includes a 20% sample of all inpatient discharges from U.S. community hospitals and is weighted to generate national estimates. Because the dataset does not include any unique patient identifiers, this study was exempt from institutional review board approval.

### Study Population

All hospitalizations from 2016 to 2020 for patients aged 20 years and older were included. Hospitalizations for systolic and diastolic heart failure were identified using the International Classification of Diseases, Tenth Revision, Clinical Modification (ICD-10-CM) codes.

Specifically, systolic heart failure (SHF) was identified using code I50.2, and diastolic heart failure (DHF) was identified using code I50.3. Demographic information such as age, sex, and race was used for stratified analyses.

### Study Outcomes

The primary outcomes analyzed were age-adjusted hospitalization rates for SHF and DHF, expressed per 100,000 population. Rates were stratified by year, sex, and race to analyze changes in hospitalization volume over the time period studied and proportional distribution by demographic subgroup.

### Statistical Analysis

All analyses were conducted using data from adults (≥20 years) hospitalized between 2016 to 2020. We conducted a retrospective analysis using data from the Healthcare Cost and Utilization Project (HCUP) National Inpatient Sample (NIS) from 2016 to 2020. The NIS is the largest publicly available all-payer inpatient healthcare database in the United States and approximates a 20% stratified sample of all U.S. community hospital discharges, allowing for weighted national estimates. Annual frequencies of SHF and DHF were identified using International Classification of Diseases, Tenth Revision (ICD-10) diagnosis codes.

To account for changes in population structure over time, age-adjusted rates per 100,000 population were computed using the direct standardization method, with the 2000 U.S. standard population as the reference. Age adjustment was performed using standard age groupings consistent with CDC guidelines. Data were analyzed using STATA 19 (Stata Corporation, College Station, TX), incorporating NIS sampling design elements, including discharge weight, to generate nationally representative estimates.

## Results

From 2016 to 2020, there were approximately 7,364,023 hospitalizations for SHF and 10,064,223 hospitalizations for DHF. The average age of the population was 58.5 years while the average age of those admitted for SHF was 69.3 years, and the average age of those admitted for DHF was 73.6 years. Women accounted for 36.2% of SHF admissions and 59.0% of DHF admissions. While Black patients made up 15.2% of the overall study population, they accounted for 19.9% of SHF hospitalizations. Medicare was the primary payer for 48.0% of the total population, but this proportion was significantly higher among those hospitalized for SHF (69.1%) and DHF (79.84%) (*Table 1*).

**Table 1:**
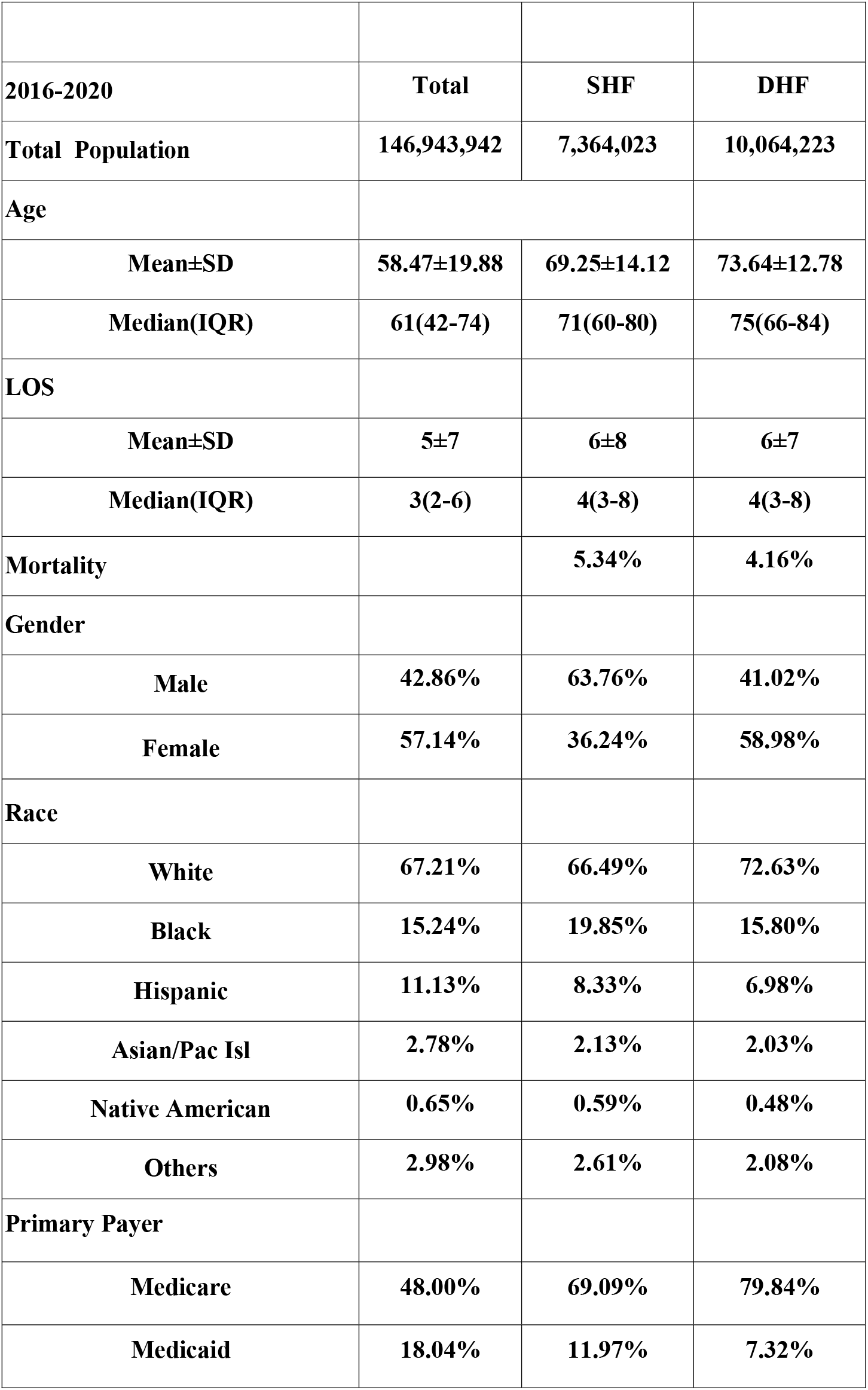
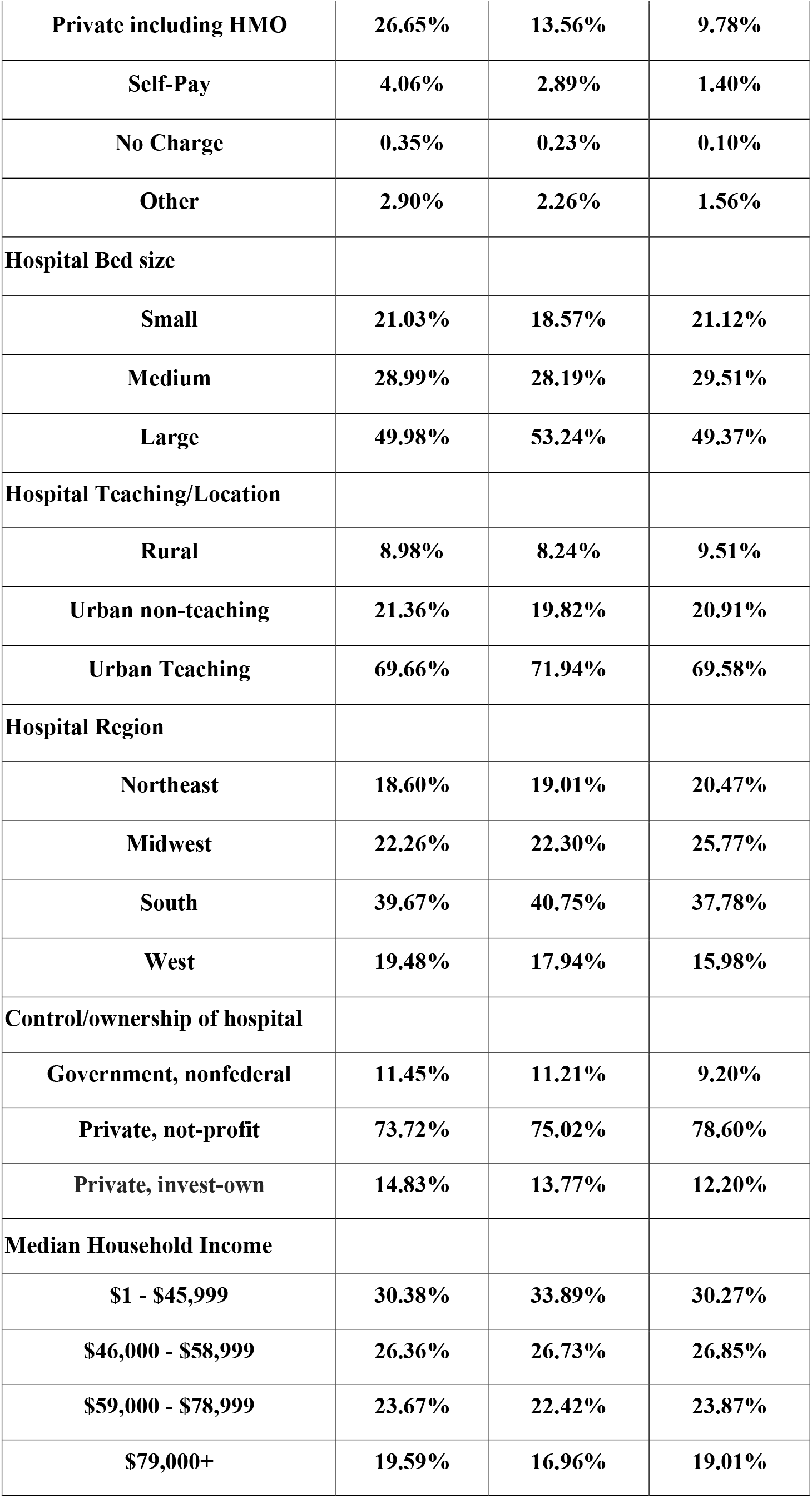
Demographics and clinical characteristics of patients admitted to the hospital for SHF or DHF.

Over the time period studied, there was an overall increase in hospitalizations for both SHF and DHF. The age-adjusted hospitalization rate for diastolic heart failure (DHF) rose steadily from 219.4 per 100,000 (95% CI: 201.4–237.1) in 2016 to 303.1(95% CI: 277.7–328.5) in 2020, representing a 38.2% increase. The systolic heart failure (SHF) hospitalization rate increased from 211.7 per 100,000 (95% CI: 194.7–228.7) in 2016 to 262.6 (95% CI: 240.6–284.6) in 2020; a 24.0% increase over the study period. The upward trend in both SHF and DHF hospitalizations was observed across all years. Additionally, the overall hospitalization rate for DHF was higher than the rate for SHF across all years, though 95% confidence intervals did overlap. (*Figure 1*).

**Figure 1:**
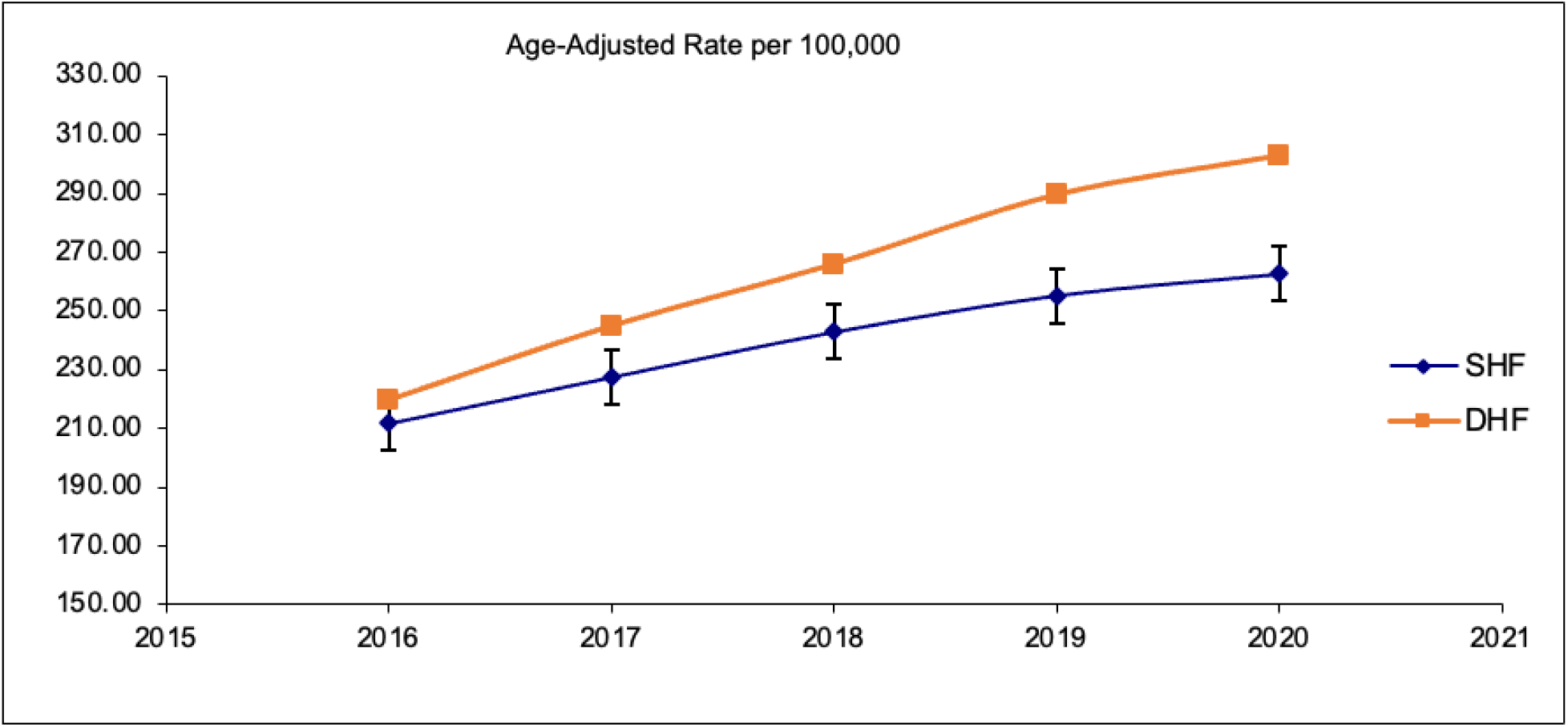
Age-adjusted rate of hospitalizations for SHF (systolic heart failure) and DHF (diastolic heart failure) from 2016-2020.

Across all years, the SHF hospitalization rate in men was significantly higher than the SHF hospitalization rate in women. In 2016, SHF rates were 302.9 per 100,000 (95% CI: 265.4– 340.4) in men and 140.3 (95% CI: 125.5–155.0) in women; by 2020, these rates had increased respectively to 370.6 (95% CI: 323.8-417.4) in men and 171.9 (95% CI: 152.6–191.1) in women (*Figure 2*). The same disparity was not observed in DHF hospitalizations. The age-adjusted DHF hospitalization rates were lower in men than in women across all years, however, 95% confidence intervals were overlapping (*Figure 3*).

**Figure 2:**
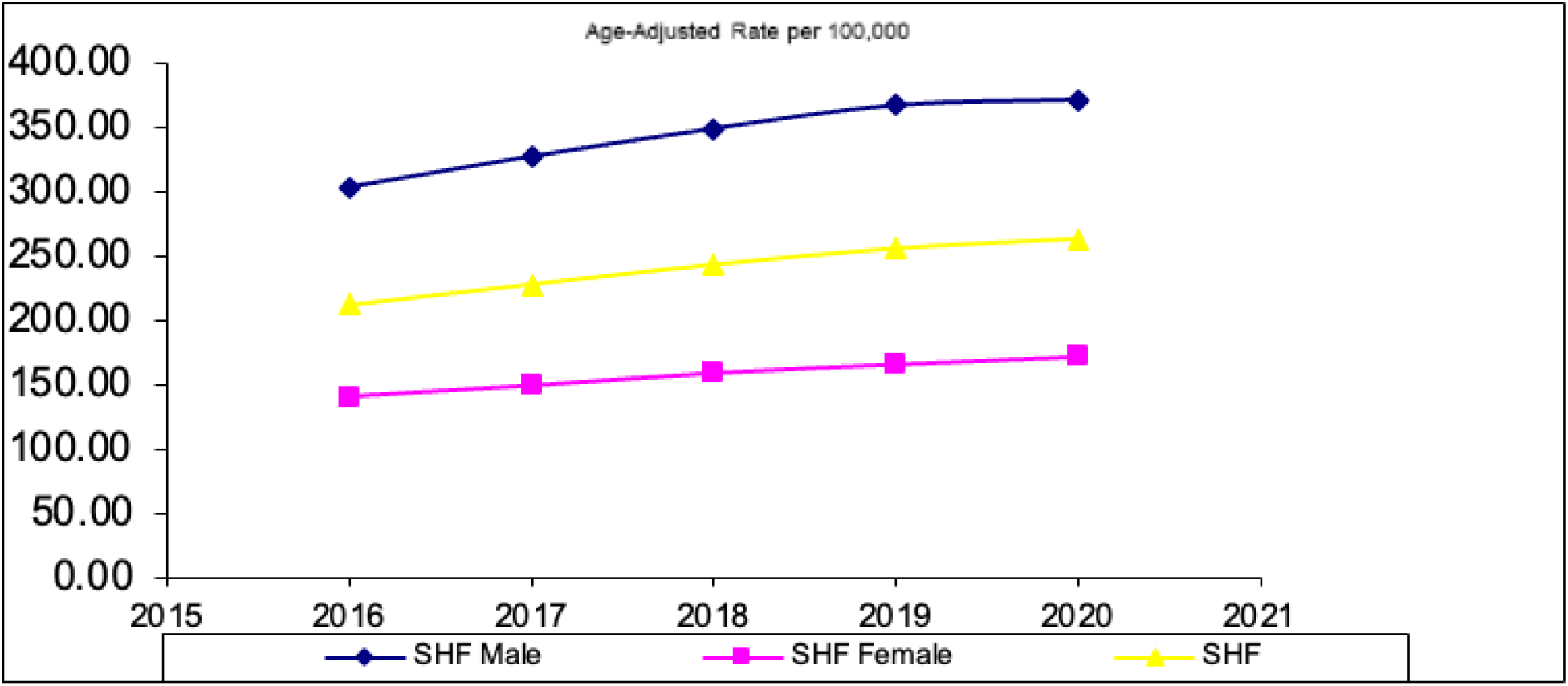
Age-adjusted rate of hospitalizations for SHF (systolic heart failure)from 2016- 2020 by sex.

**Figure 3:**
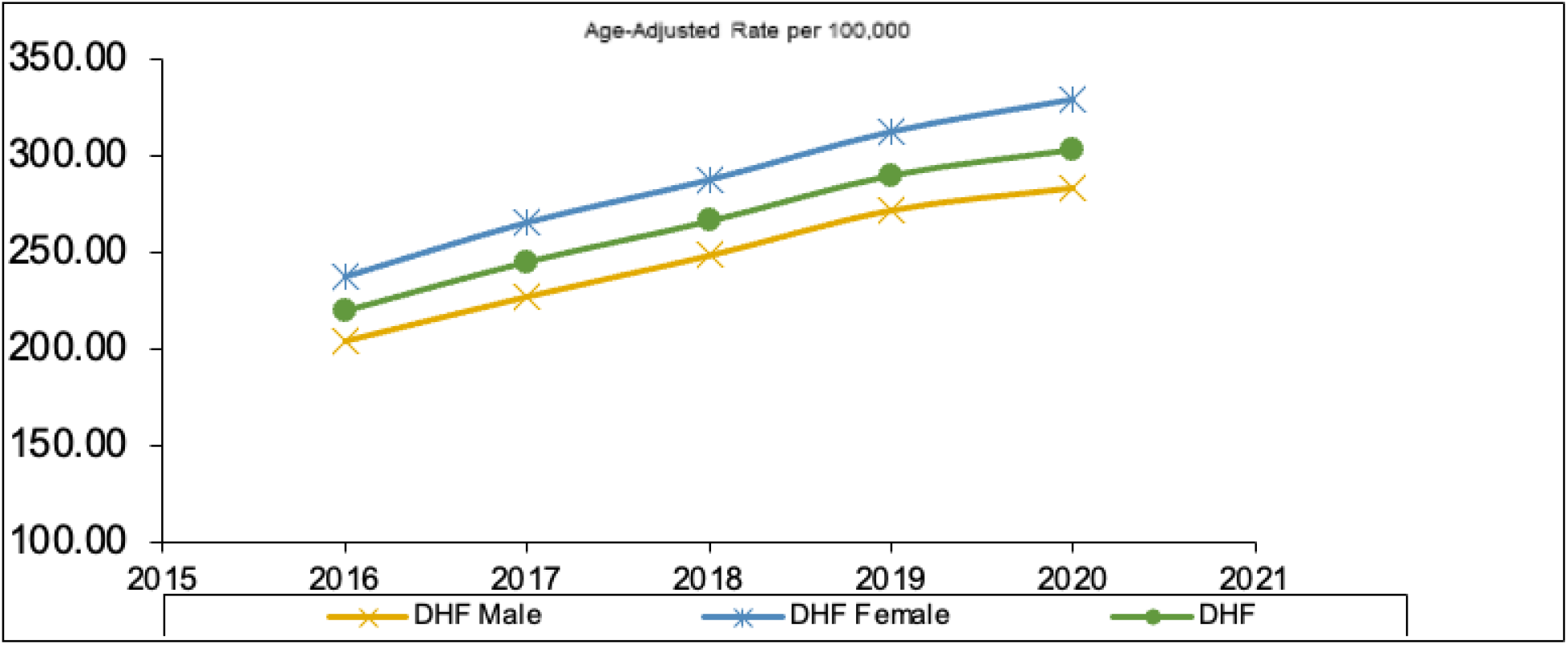
Age-adjusted rate of hospitalizations for DHF (diastolic heart failure) from 2016- 2020 by sex.

Notable racial disparities were observed in the hospitalization rates for SHF and DHF. Across all years, Black patients had the highest age-adjusted rates of hospitalization for both SHF and DHF. In 2020, SHF rates were 403.6 per 100,000 (95% CI: 317.3–489.8) among Black adults, compared to 227.5 (95% CI: 203.7–251.3) among White adults (*Figure 4*). That same year, the DHF admission rate was 418.3 per 100,000 (95% CI: 328.9–507.7) in Black adults and 284.8 (95% CI: 255.0–314.6) in White adults (*Figure 5*). Hispanic and Asian populations had modestly higher rates of SHF and DHF hospitalizations compared to Whites, however, 95% confidence intervals were overlapping (*Figures 4 and 5*).

**Figure 4:**
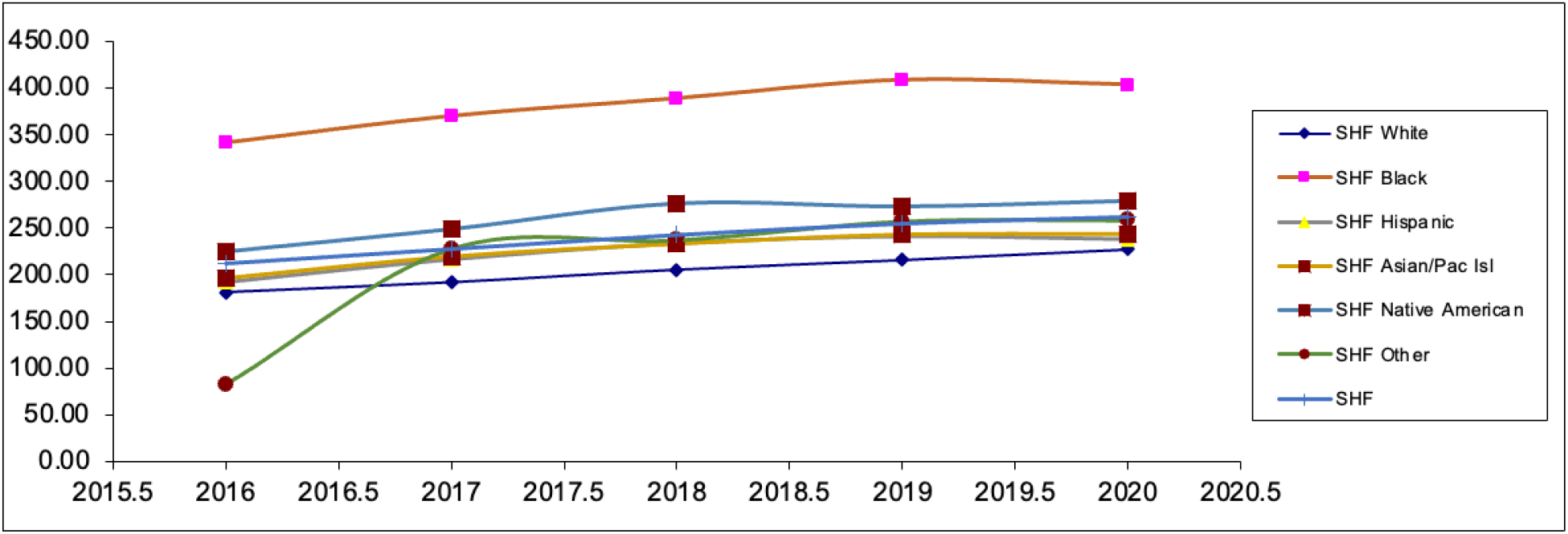
Age-adjusted rate of hospitalizations for SHF (systolic heart failure) from 2016- 2020 by race.

**Figure 5:**
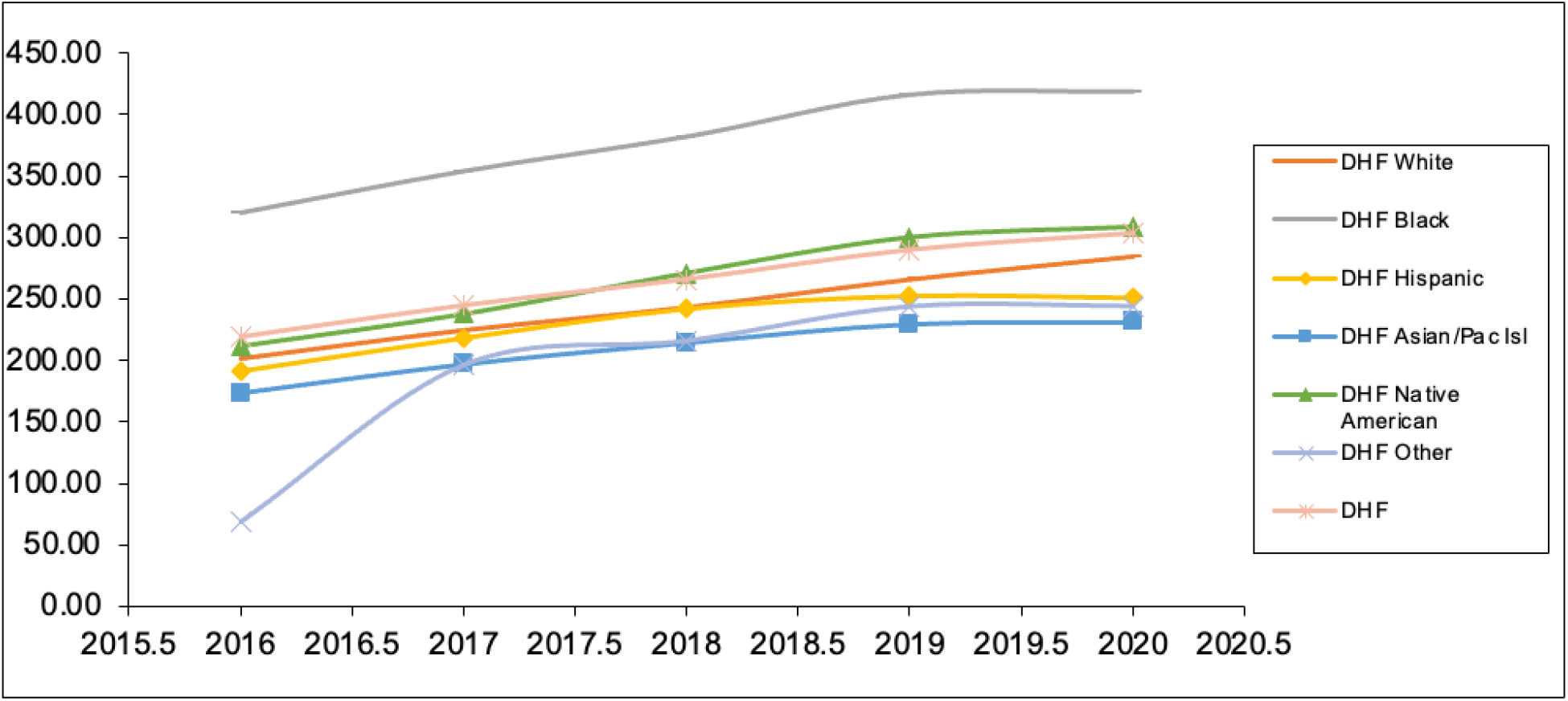
Age-adjusted rate of hospitalizations for DHF (diastolic heart failure) from 2016-2020 by race.

## Discussion

In this analysis of national age-adjusted hospitalization trends, we observed a significant increase in hospitalizations for both systolic (SHF) and diastolic heart failure (DHF) from 2016- 2020. The DHF rate increased by 38.2%, from 219.4 to 303.1 per 100,000, while SHF rose by 24.0%, from 211.7 to 262.6 per 100,000. These findings align with projections that HF prevalence is expected to rise substantially over the coming decades. ^5, 6, 7^ However, given that these projected increases have largely been attributed to the aging population, ^5, 6, 7^ it is notable that this rise in hospitalization rates was observed despite age adjustment, suggesting that factors beyond demographic shifts may be contributing.

Previous research has demonstrated declining hospitalization rates for HF overall from the early 2000s until 2013. ^18, 19^ More recent analyses of national HF hospitalizations have supported the trend of declining HF hospitalizations until 2014, at which point the trend appears to have reversed. According to these analyses, HF hospitalization rates have been increasing from 2014-2018. ^20, 21,^ Our study suggests a sustained reversal of the previously declining hospitalization rates. Multiple mechanisms likely underlie this continued increase in age-adjusted hospitalization rates for both SHF and DHF, including rising cardiovascular risk factors, ^6, 34^ improved survival post-diagnosis, ^4^ or earlier identification of HF, particularly DHF. ^35^

Our findings suggest that DHF is now more prevalent than SHF in hospitalized populations. From 2016-2020, DHF accounted for more hospitalizations (10,064,223) than SHF (7,364,023). There are several potential explanations for this finding. DHF is more common among older adults and women ^10, 29, 30, 31, 32^ and is closely associated with widely prevalent comorbid conditions, including hypertension, obesity, diabetes, and chronic kidney disease. ^27, 28, 36^ As the prevalence of these comorbidities rises, DHF incidence and diagnosis are likely to rise as well. ^34^ Increased recognition and improved diagnostic criteria for DHF in recent years may also be contributing to the trend observed due to increased detection. ^37^

Our study found that women were much more frequently affected by DHF than SHF, which is consistent with prior research on the topic. ^29, 30, 31^ Women accounted for 59.0% of DHF admissions and only 36.2% of SHF admissions. One explanation for this trend is that DHF tends to present at a later age, and women have a longer life expectancy on average. ^38^ Women also tend to present with different risk factor profiles, including greater prevalence of obesity and hypertension, ^34, 39^ conditions that have been closely associated with the development of DHF. ^40, 41^ Sex-based differences in cardiac structure/remodeling and diastolic function may also contribute to this discrepancy. ^42, 43, 44^

Conversely, men consistently had higher SHF hospitalization rates than women. Men made up 42.7% of the population studied but accounted for 63.8% of SHF hospitalizations. This trend has been previously observed in the literature. ^16, 45^ Like with the sex-based discrepancies seen in DHF, it has been proposed that differences in risk factors and comorbidities underlie this disparity. Compared to women, men with HF have higher rates of underlying coronary artery disease, ^39^ which is more associated with the development of SHF than DHF. ^46^

Pronounced racial disparities were also observed. Black adults experienced significantly higher hospitalization rates than White adults for both SHF and DHF across all years. In 2020, the SHF rate among Black adults was 403.6 per 100,000, compared to 227.5 in White adults. These findings align with prior research on racial inequities in HF hospitalizations. ^13, 17,18^ Several factors may contribute to these persistent disparities, including a higher burden of comorbidities among Black adults, unequal access to specialized care, disparities in insurance coverage, lower socioeconomic status, and structural inequities within the healthcare system. ^17, 47, 48^

Overall, these findings highlight the need to develop HF subtype-specific, sex-specific, and race-conscious strategies in HF prevention, diagnosis, and management. The disproportionate burden borne by men and Black adults underscores the necessity of targeted interventions and healthcare system reforms to address long-standing cardiovascular disparities.

## Conclusion

Heart failure hospitalization rates for both SHF and DHF are increasing in the U.S. population. Between 2016 and 2020, age-adjusted hospitalization rates increased substantially for both SHF and DHF. The age-adjusted hospitalization rate for diastolic heart failure (DHF) rose by 38.2% while systolic heart failure (SHF) hospitalization rate increased by 24.0% over the study period. SHF hospitalizations were consistently more frequent among men, and hospitalizations for both HF subtypes were disproportionately higher among Black adults throughout the study period. The sex- and race-based disparities observed underscore the need for public health strategies and clinical interventions to combat the growing burden of heart failure and promote equity in cardiovascular outcomes.

## Limitations

This study has several limitations. First, the analysis was based on data regarding HF subtypes identified through ICD-10 codes, which may be affected by coding errors or misclassification.

Second, as the NIS dataset captures hospitalizations rather than individual patients, repeat admissions by the same patient may be counted multiple times. Additionally, some racial and ethnic categories, particularly Asian and Native American populations, had small sample sizes, limiting the precision of rate estimates and comparisons.

## Data Availability

NIS data base is publicialy available uppon purchase

## Notes

Conflict of interest: The authors report no financial relationships or conflicts of interest regarding the content herein

### Competing Interest Statement

The authors have declared no competing interest.

### Funding Statement

None

### Author Declarations

NIS database is publicly available without a patients identifier exempt from IRB

